# Summer School Holidays and the Growth Rate in Sars-CoV-2 Infections Across German Districts

**DOI:** 10.1101/2020.10.11.20210773

**Authors:** Thomas Plümper, Eric Neumayer

## Abstract

**Objectives:** To estimate the effect that summer school holidays had on the growth rate in Sars-CoV-2 infections across German districts. The Robert-Koch-Institute reports that during the summer holiday period a foreign country is stated as the most likely place of infection for an average of 27 and a maximum of 49 percent of new Sars-CoV-2 infections in Germany. Yet, infection may have taken place elsewhere, not all international travel is holiday-related and any impact of holiday-related travel will not be restricted to holidays abroad.

**Design:** Cross-sectional study on observational data. In Germany, summer school holidays are coordinated between states and spread out over 13 weeks. We analyse the association between these holidays and the weekly infection growth rate in SARS-CoV-2 infections across 401 German districts. Employing a dynamic model with district fixed effects, we test whether the holiday season results in a statistically significantly higher infection growth rate than the period of two weeks before holidays start, our presumed counterfactual.

**Results:** We find effects of the holiday period equal in size to almost 50 percent of the average district growth rate in new infections in Germany during their respective final week of holidays and the two weeks after holidays end. States in the West of Germany tend to experience stronger effects than those in the East. This is consistent with another result, namely that we find statistically significant interaction effects of school holidays with per capita taxable income and the share of foreign residents in a district’s population, with both factors hypothesised to increase holiday-related travels.

**Conclusions:** Our results suggest that changed behaviour during the holiday season accelerated the pandemic and made it considerably more difficult for public health authorities to contain the spread of the virus by means of contact tracing. Governments did not prepare adequately or timely for this acceleration.

## INTRODUCTION

Holidays can be expected to accelerate the Sars-CoV-2 pandemic. To a small extent, this is because travelling via bus, train or plane adds to the risk of becoming infected.^1,2,3,4^ More importantly, infections rise because individuals change their social behaviour during holidays.^5^ Many holiday-makers have more and more intense social interactions, often to people that they do not know and do not share social capital with which has been found to be conducive to maintaining social distancing norms.^6,7^ Mobility also reduces the health agencies’ ability to successfully trace close contacts of people that are infected with Sars-CoV-2.^8^

The Robert-Koch-Institute (RKI) reports that over Germany’s entire summer school holiday period – shifted forward by two weeks to allow time for a return from holidays, the incubation period and delays in becoming tested – on average in about 27 percent of weekly cases reported to the Institute a foreign country was mentioned as the most likely place of infection with a maximum of 49 percent of weekly cases reported in week 34 which is in mid-August.^9^ It is, however, problematic to interpret these numbers as the effect of holiday-related travel since some of the infections may not actually have occurred abroad, not all international travel is necessarily holiday-related and these numbers fail to take into account the impact of holidaying within Germany. These numbers may thus over-state or under-state the true impact of the holiday season on the pandemic.

We therefore broaden the perspective and analyse the extent to which summer school holidays have accelerated the pandemic in Germany, including but not limited to holiday-related travel abroad. Whilst the media debate in Germany focused on holiday-makers returning from a vacation spent abroad, it is likely that the spread of the virus would have accelerated even if all holiday-makers had stayed in Germany. The risks of holidaying are not limited to international holiday travel.

In order to estimate the effect of summer school holidays on the growth in infections, we employ an ecological analysis of variation in infection growth rates across German districts – there are no data at the individual level. Germany provides an excellent case study since we can exploit a particular feature of the system of school holidays in this country, namely that they are not uniform across the Federal Republic but vary in their start and therefore also their end date from state to state in a pre-determined way. This idiosyncratic feature allows us to disentangle the effect that holidays have on new infections in German districts located in states that are or have been on holiday from the general upward trend in new infections in Germany.

There is surprisingly little existing evidence on the impact of public holidays on the Sars-CoV-2 pandemic. Some judge the extension of the Lunar New Year holidays in China as positively contributing to the country’s successful containment of the pandemic but this represents a rather unique setting given the pandemic only started around the time of this holiday period and the holiday extension helped authorities to identify infected individuals before travelling home.^10,11^ Others find that Israel’s hitherto successful mitigation policy broke down in the wake of mass social gatherings during the 9 to 11 March Jewish holiday of Purim or simulate which measures may increase or decrease the impact of holiday-related travel from metropolitan areas to the provinces in Sweden on infections.^12,13^ To the best of our knowledge, our is the first academic study of the impact that the summer school holiday season has actually had on the pandemic.

## BACKGROUND

The most straightforward argument against the hypothesis that holiday travels to foreign places increase infections back home is simple: if a holiday-maker travels to a destination that has fewer active cases per capita than her home area, the probability of an infection declines, all other things equal. Yet, not only did Germany have lower infection rates than many foreign holiday destinations. More importantly, all other things are not equal and, for at least three reasons, we would expect that holiday-related travels increase infections regardless of the relative difference in infection rates between Germany and other countries.

First, the process of travelling may in itself increase the risk of an infection. If the traveller journeys to an airport, train or bus station, waits around for boarding and then boards a plane, train or bus, a potentially crowded indoor space with imperfect ventilation, the probability of transmission through aerosols increases – small exhaled droplets that may contain the virus. The evidence on infections during flights and train rides is rather mixed. In unpublished work, Hu et al. report that the probability of an infection while sitting within 3 rows and 5 columns from an infected passenger is low on average (0.32 percent) but can be as high as 10.3 percent.^3^ Likewise, Schwartz et. al. find no contagion on a (single) flight with infected passengers. They suggest that “studies of airplane transmission are commonly biased by contacts sharing exposure risks before boarding the air-craft.”^4^ However, the absence of evidence of an effect is not evidence of absence of such an effect. As Gonne and Hubert argue, travel restrictions are surprisingly easy to implement, but their efficacy is very difficult to assess.^2^ Moreover, though very few infections have been demonstrably associated with trains and planes, there is at least some evidence that public transportation has contributed to the spread of the virus in the early stages of the pandemic in New York City.^14,15^ All in all, however, we would presume this first factor to be a relatively minor one relative to the other two.

Second, people who go on vacation do not follow the same daily routines as they do at home. They are less likely to follow social distancing norms, partly because they follow holiday routines and partly because social control from family and friends declines. Migrant workers visiting their home countries will find it hard to follow rules of social distancing when visiting friends and family with long-established social courtesy norms are often stronger than relatively new and untrained social distancing rules, partly because of the misperception that family and friends do not pose a serious risk. For migrant workers a summer holiday usually implies family visits, exchange of gifts, and parties. Likewise, tourists spend more time on leisure activities and visit bars and restaurants more often than they would at home. In an interview with the *Time* magazine on 27 August 2020, Martin McKee, professor of public health at the London School of Hygiene and Tropical Medicine, told journalist Madelaine Roach that the rise in new infections was “most likely” triggered by “tourists in crowded bars, restaurants, and nightclubs.” There exists plenty of video and photo footage that shows tourists violating social distancing norms. What appears to be lacking, however, is systematic research on the behavioural-related differences of the same people at home and during their holidays as it could be that the same people shown on the footage also violate social distancing norms back home. In a computational model on H1N1 influenza, Shi et al. find that social gatherings and travels increase infections close to the peak of a pandemic, but have little effect when the number of active infections per capita is low.^16^ While this finding may be plausible for influenza where virtually all contagious people show symptoms, it does not need to hold for a virus that can be transmitted from asymptomatic infected people as is the case with Sars-CoV-2. We therefore find it plausible that holidays reduce the willingness and ability of people to adhere to social distancing norms.

Third, holiday travellers have more social interactions with people that they do not normally physically interact with and/or they do not know. Holiday travel inevitably implies interacting with more strangers or faintly acquainted individuals than back home. As a consequence, infected people will usually not be able to name their recent close contacts after they have been diagnosed with the virus. What makes matters worse is that tracing apps are incompatible across European countries. Hence, with holiday travel-related transmissions, contact tracing becomes prohibitively difficult: travellers are unlikely to identify many of their close contacts and technology will also be of little help since contact tracing apps do not work across European borders. Thus, even if the behaviour of holiday travellers were not to increase the spread of Sars-CoV-2 as such, the declining ability to stop the spread through contact tracing measures should eventually lead to an increase in infections.

In sum, holiday-related travels increase infections not necessarily because of the act of travelling itself though this might also contribute. Rather, travelling increases infections because tourism and family visits abroad have a profound effect on the number, intensity and nature of social interactions and significantly reduce the ability to trace social contacts. This logic implies that the odds of catching an infectious disease may increase even if travellers’ spend their vacation in a region of their home country or another country with a lower infection rate. Likewise, though, the risk becomes larger the higher the rate of infection in the traveller’s destination, if travellers do not adjust their behaviour to the higher risk.

## METHODS

Our dependent variable is the weekly percentage growth rate in the number of infections in a German district. Infection data by district are sourced from the RKI website (www.rki.de). We also employ two explanatory variables that we model as conditioning the effect of summer school holidays, namely a district’s average taxable income in tens of thousands of Euros and the fractional share of foreigners amongst a district’s resident population with data sourced from the regional database of the German statistical office (www.destatis.de). Due to lack of disaggregated data for these two variables, the twelve districts of Berlin are aggregated to one single city state district. Our sample then covers all 401 districts in Germany with the temporal dimension drawn from the period starting with the weekly growth rate to Wednesday 10 June (week 23) and terminating with the weekly growth rate to Wednesday 23 September (week 38). We deliberately define the week to end in a Wednesday rather than Sunday or Monday to avoid noise from occasional corrections made on Mondays or Tuesdays to compensate for under-reporting over the weekend. For each district, we analyse the period ranging from two weeks prior to the beginning of holidays to two weeks after the end of the holidays. Our panel thus has N=401 districts and T=10 weeks equals 4010 observations.

In Germany, the dates of the summer school holidays are chosen years in advance by each of the 16 states in close consultation with each other. The intention is to reduce the probability and length of traffic jams on Germany’s crowded motorways during the summer months. In each state, schools close for approximately six weeks. In 2020, the summer school holiday season began on June 22 in Mecklenburg-Vorpommern and ended September 9 in Baden-Württemberg. Hence, Germany spreads the holiday season over almost 13 weeks.

The average weekly growth rate of infections across all German districts over the entire sample period is 2.70 percent with a standard deviation of 3.48 percent. In Germany as a whole, the number of new infections had been stable at around 500 per day until the end of July. In August, the number of daily infections begun to rise reaching approximately 1,500 new daily infections at the end of August and about 2,000 daily new infections at the end of September. This upward trajectory in infections in Germany coincides with the summer school holiday season. It is unlikely, however, that the return of rising infection numbers has been determined by school holidays alone. To isolate the predicted effects from other influences, we include a lagged dependent variable in the model that accounts for the common trend in the data.^17,18^ Results are similar if we use an alternative approach for taking out the common trend, such as an autoregressive model. This is a conservative research strategy since part of this trend was most likely caused by returning holiday-makers. However, it is impossible to provide a precise estimate of the influence of holidays on the common trend because holiday travel was allowed in all states and all districts at all times, not just during school holidays.

Most of our estimation models are based on a specification with a dummy variable that is set to 1 if a district is located in a state in which schools are on summer holidays in that week as well as a dummy variable for the two-week period after the holidays. This specification can be interpreted as a Chow-type model in which the dummy variables estimate whether there is a structural break between the holiday period as well as the period of two weeks after the holidays end, both relative to the period of two weeks before holidays begin, the presumed counterfactual.^19,20^ We include a dummy variable for the period of two weeks after holidays officially end to capture the impact caused by late returners from holidays and to account for the incubation time which is estimated to be up to 2 weeks in some cases as well as potential delay in becoming tested. This relatively simple specification with two dummy variables only is handy for extensions where we allow their effect to vary by state and allow their effect to be conditioned by two district-level variables that are likely to impact on the number of holiday-related travels undertaken from each district (on which more below). It is not an optimal specification however given it presumes the effect to be constant within the holiday period. Empirical evidence suggests that the average length of holiday stay of German tourists is about 12-14 days.^21^ Therefore, one should expect that infections should start to rise only about 2 to 4 weeks after the beginning of the school holidays. Therefore, we will also present results from a more appropriately specified model that allows the effect of the holiday period to vary week-by-week.

We include district fixed effects to absorb any variation across districts that is time-invariant such as demographic, geographic and socio-economic factors that render some districts more generally exposed to the pandemic than others.^22^ Since potential control variables come from annual data, they are time-invariant for the specific panel structure we have. These time-invariant variables are perfectly collinear with the district fixed effects and we therefore cannot estimate their effect in a district fixed effects model. They can however condition the effect of the time-varying school holidays variables, as we do in one model employing average taxable income and the share of foreigners amongst a district’s resident population as conditioning variables. We cluster standard errors on districts. If we additionally apply two-way clustering of standard errors also by states results are hardly affected (results not reported).

We test three hypotheses. First, school holidays have a positive effect on the infection growth rate in a district. Second, the later parts of any given holiday season should have a larger effect than its earlier parts given that there is a delay until holiday travellers return home and given infections are on the rise in practically all holiday travel destinations, both within and outside Germany, thus increasing the risk of catching the virus as the holiday season proceeds. Third, school holidays will have a stronger effect on the infection growth rate in districts that are richer on average and in which a larger share of the resident population are foreigners. Richer people can afford better to go on holiday for longer and foreign citizens are likely to use the holiday season for returning to their home country for family visits (possibly in addition to taking other holidays) not least because the lockdown in the spring prevented most of them from seeing family abroad over the Easter holiday period.^23^

## RESULTS

In table 1 we first of all report results on a dummy variable that is set to 1 if a district is located in a state in which schools are on summer holidays in that week as well as a dummy variable for the two-week period after the holidays (model 1). We find that the summer school holiday weeks are on average predicted to increase the infection growth rate by 0.72 (95% CI 0.41-1.03) percentage points relative to the period before holidays, consistent with our first hypothesis. The two-week period after holidays end is predicted to increase the weekly infection rate by 1.96 (95% CI 1.56-2.35) percentage points.

**Table 1.**
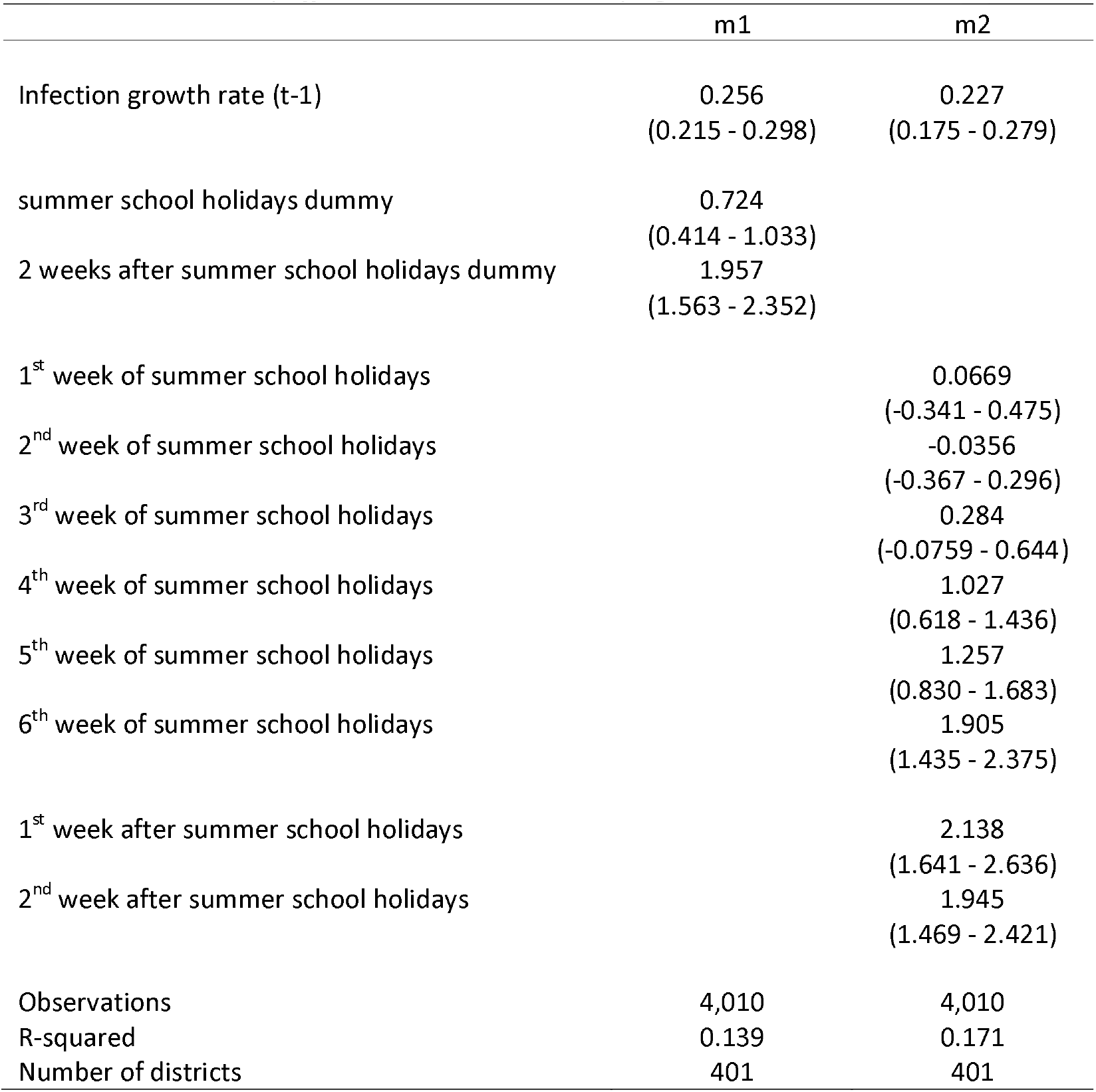
School Holiday Effects, Pooled and Time-varying

|  | m1 | m2 |
| --- | --- | --- |
| Infection growth rate (t-1) | 0.256<br>(0.215 - 0.298) | 0.227<br>(0.175 - 0.279) |
| summer school holidays dummy | 0.724<br>(0.414 - 1.033) |  |
| 2 weeks after summer school holidays dummy | 1.957<br>(1.563 - 2.352) |  |
| 1<sup>st</sup> week of summer school holidays |  | 0.0669<br>(-0.341 - 0.475) |
| 2<sup>nd</sup> week of summer school holidays |  | -0.0356<br>(-0.367 - 0.296) |
| 3<sup>rd</sup> week of summer school holidays |  | 0.284<br>(-0.0759 - 0.644) |
| 4<sup>th</sup> week of summer school holidays |  | 1.027<br>(0.618 - 1.436) |
| 5<sup>th</sup> week of summer school holidays |  | 1.257<br>(0.830 - 1.683) |
| 6<sup>th</sup> week of summer school holidays |  | 1.905<br>(1.435 - 2.375) |
| 1<sup>st</sup> week after summer school holidays |  | 2.138<br>(1.641 - 2.636) |
| 2<sup>nd</sup> week after summer school holidays |  | 1.945<br>(1.469 - 2.421) |
| Observations | 4,010 | 4,010 |
| R-squared | 0.139 | 0.171 |
| Number of districts | 401 | 401 |

Model 1, which pools all holiday weeks together, masks that the effect is likely to vary and to increase over the holiday period. Model 2 is more appropriately specified as it allows the effect of the holiday season to vary week-by-week. We find that the effect increases in later weeks of the school holidays. The effect is essentially zero in the first two weeks, rises from week three onwards, becomes statistically significant from week four onwards and then increases to 1.91 (95% CI 1.44-2.38) percentage points in week 6.

The coefficients of the first and the second week after school holidays finish show that the increases in infection growth brought about by the school holidays do not disappear but essentially remain the same as in week 6. In terms of substantive importance of this finding, 1.91 (95% CI 1.44-2.38) percentage points toward the end of the holiday season equates to 48.7 (95% CI 36.6-60.6) percent of the average growth rate across German districts during their sixth week of holidays, which is higher than the average growth rate during the entire sample period and therefore represents the more appropriate benchmark against which the substantive effect size should be assessed so as not to over-state it. For the first and second week after holidays, the equivalent computation would suggest effects that equate to 49.5 (95% CI 40.0-61.0) and 46.1 (95% CI 34.8-57.4) of the average weekly growth rate in those weeks.

In model 3, reported in table 2, we allow the structural breaks to vary state-by-state but revert back to the simple Chow-type structural break model with only two dummy variables per state as otherwise we would have to report or visualize well over a hundred coefficients. We exclude the two states of Hamburg and Berlin since both are counted as consisting of only one district in our data, which would result in unreliable estimates in a district fixed effects specification. Table 2, in which we sort states by the point estimate of the holiday period dummy variable, shows large variation in the holiday effect on infection growth rates across districts in different German states. Overall, we find that richer states are more likely to show relatively large effects, and we find that the increase in infections associated with the holiday season tends to be larger in the Western German states than in the Eastern German states. Looking state by state, we find a statistically significant effect of the holiday period or the two-week period after the end of holidays or of both in 10 of the 14 states included in model 3.

**Table 2.** School Holiday Effects, Varying by State

| m3 | during school holidays | after school holidays |
| --- | --- | --- |
| Infection growth rate (t-1) | 0.246<br>(0.202 - 0.290) |  |
| Lower Saxony | 2.033<br>(1.392 - 2.674) | 0.200<br>(0.135 - 0.266) |
| Baden-Württemberg | 1.178<br>(0.743 - 1.613) | 1.536<br>(1.039 - 2.033) |
| Bavaria | 1.118<br>(0.835 - 1.401) | 1.213<br>(0.932 - 1.493) |
| Schleswig-Holstein | 1.116<br>(0.672 - 1.559) | 0.168<br>(0.101 - 0.234) |
| Bremen | 0.938<br>(0.458 - 1.418) | 0.339<br>(0.0234 - 0.654) |
| Hesse | 0.845<br>(0.230 - 1.460) | 0.571<br>(0.396 - 0.746) |
| Saxony | 0.799<br>(0.379 - 1.220) | 0.209<br>(0.0793 - 0.338) |
| Saxony-Anhalt | 0.703<br>(-0.233 - 1.638) | 0.0300<br>(-0.0327 - 0.0927) |
| Thüringen | 0.593<br>(-0.0584 - 1.245) | 0.0591<br>(-0.00511 - 0.123) |
| Rhineland-Palatinate | 0.329<br>(-0.331 - 0.990) | 0.255<br>(0.170 - 0.341) |
| Saarland | 0.171<br>(-0.612 - 0.954) | 0.0828<br>(0.00638 - 0.159) |
| Mecklenburg-Vorpommern | -0.290<br>(-1.614 - 1.034) | 0.381<br>(0.0665 - 0.696) |
| North Rhine-Westphalia | -0.352<br>(-2.222 - 1.519) | 0.137<br>(-0.0603 - 0.334) |
| Brandenburg | -1.123<br>(-2.389 - 0.144) | 0.0241<br>(-0.416 - 0.464) |
| Observations | 4,010 |  |
| R-squared | 401 |  |
| Number of districts | 0.163 |  |

Only districts in Thüringen, Saxony-Anhalt and Brandenburg, all located in the East, and North Rhine-Westphalia show no statistically significant effects. The three Eastern German states have low and slowly but linearly rising growth rates in infection numbers and therefore exhibit no significant holiday effects. In contrast, North Rhine-Westphalia had high infection rates and growth rates above 4 percent already before the holidays begun due to super-spreader events in a slaughterhouse of the Tönnies company in the districts of Gütersloh and Warendorf with no further acceleration during the holiday period. These events have reduced the estimate of holiday-related growth in infections for this state because the very strong decline of new infections in these two districts after containment measures were put in place locally compensates for the holiday-related increase in the growth rate of infections in all other districts of North Rhine-Westphalia. In fact, if we drop the two districts of Gütersloh and Warendorf from the estimations then both coefficients of the holiday and post-holiday periods become statistically significantly positive for this state.

Figures 1a and 1b show cumulative infection numbers (blue line – left scale) and the infection growth rate with respect to the past seven days (bars – right scale) for each day between day 167 and day 267 of 2020 for two states, namely Bavaria, the richest German state bar the two city states of Bremen and Hamburg, and Saxony-Anhalt, the state with the lowest average per capita income. As Model 3 has shown, the holiday season was associated with a significant increase in infections in Bavaria, while we did not find a significant increase in infections relative to the trend in Saxony-Anhalt. Figures 1a and 1b support and illustrate these findings from our regression analysis. The boundaries of the grey-shaded area indicate the first and the last day of school holidays. Even without econometric controls for the common trend in the data, one can see a structural break in Bavaria’s infection dynamics but no such structural break for Saxony-Anhalt.

**Figure 1a:**
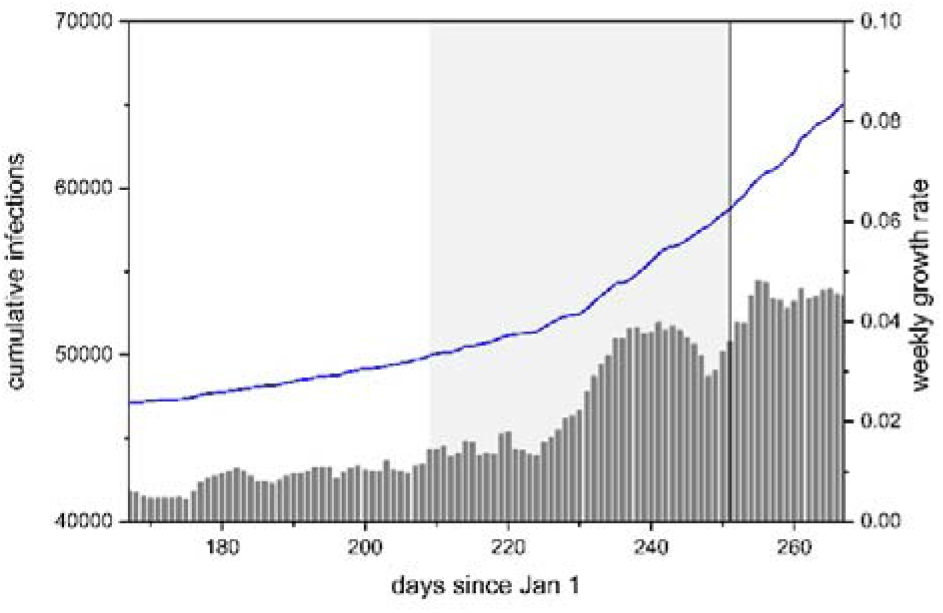
School Holidays and Infections in Bavaria

**Figure 1b:**
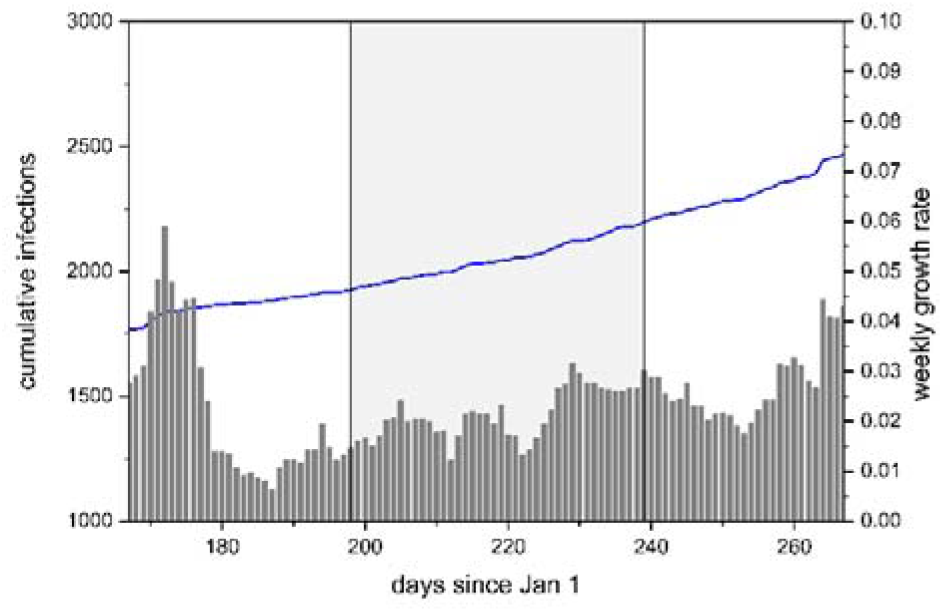
School Holidays and Infections in Saxony-Anhalt

Bavaria’s growth rate in infections is slowly increasing from about 0.4 percent on day 167 to about 1 percent on the day its summer holidays started. Two weeks later, when holiday-makers began to return, the growth rate still stood at 1.06 percent (day 226), but quickly rose to 3.87 percent on day 237. We find no such effect for Saxony-Anhalt. This state had a short outbreak of infections in mid-June, with the infection growth rate afterwards first falling to below 1 percent and then starting to climb almost linearly over the summer, with no obvious structural break triggered by the beginning of the school holidays.

Saxony-Anhalt and Bavaria differ in many respects. Bavaria is richer, more industrial, more urbanized, and it also hosts a larger share of foreign residents. In table 3, we allow the effect of summer school holidays and the two-week period after holidays to be conditioned by one or both of two variables, namely by average taxable income in a district as well as by the share of foreigners amongst a district’s residents. These variables are time-invariant for our sample, therefore we cannot estimate coefficients for these variables themselves in a model with district fixed effects. However, we can estimate the conditioning effect of these variables on the time-varying holiday variables.

**Table 3.** The Conditioning Effect of Average Taxable Income and of the Share of Foreign Residents

|  | m4 | m5 | m6 |
| --- | --- | --- | --- |
| infection growth rate (t-1) | 0.255<br>(0.213 - 0.296) | 0.251<br>(0.209 - 0.294) | 0.251<br>(0.208 - 0.293) |
| Summer school holidays dummy | -0.669<br>(-1.794 - 0.456) | 0.391<br>(-0.0131 - 0.796) | -0.627<br>(-1.735 - 0.480) |
| post-holidays dummy | 0.107<br>(0.0223 - 0.191) |  | 0.0335<br>(-0.0463 - 0.113) |
| holidays dummy*tax. income p.c. | 0.0770<br>(0.0126 - 0.141) |  | 0.0664<br>(0.00400 - 0.129) |
| post-holidays dummy*tax. income p.c. | 0.0333<br>(-1.483 - 1.550) | 0.836<br>(0.288 - 1.383) | 0.322<br>(-1.147 - 1.791) |
| holidays dummy*share foreign residents |  | 3.031<br>(-0.467 - 6.530) | 1.393<br>(-1.824 - 4.609) |
| post-holidays dummy*share foreign resid. |  | 10.23<br>(5.489 - 14.97) | 9.405<br>(4.856 - 13.95) |
| Observations | 4,010 | 4,010 | 4,010 |
| Number of districts | 401 | 401 | 401 |

Model 4, reported in table 3, shows a positive and statistically significant interaction effect between average taxable income and the dummy variables for school holidays and for the two-week post-school holiday period. Model 5, also reported in table 3, shows a positive and statistically significant interaction effect between the share of foreigners amongst a district’s residents and the post-holiday period dummy. The two conditioning variables are correlated at r = 0.44 with each other. In model 6 we include interaction effects with both taxable income per capita and the share of foreigners amongst a district’s residents. The conditioning effects are rather similar in model 6 as they were in models 4 and 5, except the one between the post-holiday dummy variable and taxable income per capita is no longer statistically significant. In substantive terms, the results from model 6 imply that the effect of the holiday period is almost eleven times stronger in the richest than in the poorest districts (1.73 as opposed to 0.16 percentage point increase), while the effect of the two-week post-holiday period is almost ten times stronger in districts with the highest share of foreign residents than those with the lowest share (4.25 as opposed to 0.44 percentage point increase).

## DISCUSSION

We have analysed the association between summer school holidays and the weekly growth rate of Sars-CoV-2 infections in German districts. We have argued that the holiday season increases infections because both domestic and international vacations as well as family visits abroad have a profound effect on the number, intensity and nature of social interactions and significantly reduce the ability of public health authorities to trace close contacts of infected holiday-makers.

While infection numbers probably would have gone up to some extent in the counterfactual case of no summer holidays because large parts of the population have become tired of social distancing, we have found a substantively important increase in the growth rate of infections associated with the holiday season. Disaggregating the effect week-by-week, we find that the effect increases over the holiday period, becoming statistically significant from week 4 onwards and not reverting back to the growth rate from before the holiday period in the two weeks after holidays end. We have found that by the end of the holiday period the estimated effect equates to 48.7 (95% CI 36.6-60.6) percent of the average growth rate across German districts during their respective final week of holidays and to 49.5 (95% CI 40.0-61.0) and 46.1 (95% CI 34.8-57.4) of the average growth rate during their respective first two weeks after holidays end. Despite being based on a research design that captures the effect of holidaying both within and outside Germany, our central estimates are only slightly higher than the maximum of cases reported to the RKI during Germany’s holiday season for which a country abroad is stated as the most likely place of infection, which reach a maximum of around 49 percent in week 35 in mid-August with close to 45 percent in the two weeks either side of this maximum. It is interesting to see that two very different approaches produce quite similar estimates.

Beyond reporting evidence in support of the claim that holidays increase infections on average, we have explored how the effect differs across German states and have shown that there are statistically significant structural breaks in the growth rate of infections in at least 10 of the 14 German states with more than one district in our dataset. The stronger effects take place in the Western German states. In fact, those without a statistically significant structural break are all located in Eastern Germany with the exception of North Rhine-Westphalia which is however an outlier due to a very large local outbreak before the holiday season. Two main hypothesised reasons for this heterogeneity across German states were that the states with a stronger effect consist of districts that tend to be both richer and have a larger share of foreign residents amongst their population, both of which spurs holiday-related travel. Corroborating this, we have shown that the higher is per capita income and the higher the share of foreigners in a district, the larger the increases in the growth rate of infections. This would result in higher than average effects for Hamburg, Baden-Württemberg, Bavaria and Hesse, all of which are dominated by districts with above average income and an above average resident share of foreigners, and lower than average effects for each single state in Eastern Germany, with the exception of Berlin, which are all relatively poor (by German standards) and have a low share of foreigners amongst their residents.

There are important limitations to our research design. First, there are the well-known limitations of any ecological study like ours. Ideally, one would employ individual-rather than district-level data. We had to resort to district-level data since data on individuals across Germany do not exist. In an ideal world, one would be able to trace back infections to individual behaviours during holidays. However, the necessary data do not exist and – due to privacy protection policies – cannot be collected. Second, we can only capture the effect of holiday-related travels triggered by public summer school holidays. Families with children of school-age in particular are dependent on school holidays for their holiday travel and the same holds for the employees of firms that close down for company holidays over the summer school holiday period. Thus, the majority of holiday travels will take place during school holidays. Yet, not all of holiday-related travel takes place during school holidays, which potentially biases downwards our estimate of the effect of holiday-making on Sars-CoV-2 infection.

## CONCLUSION

Summer school holidays have substantially contributed to the rise in infections in Germany. These holiday effects were entirely predictable and yet public health authorities largely failed to mitigate the impact. Of course, notwithstanding the fact that Germany like other countries imposed travel restrictions with scant regard to international health regulations in the spring of 2020,^24^ it would not have been realistic for either the German federal government or state governments to implement let alone enforce prohibitions on holiday-related travel over the summer. However, neither the federal nor the state governments systematically kept reminding travellers of the importance of maintaining social distancing and keeping track of close contacts. Doing so may have been futile anyway but they did not prepare to deal with the predictable rapid increase in infections either. What they could and arguably should have done was to significantly drive up testing facilities to compensate for the increase in infections and the reduced contact tracing capabilities. Eventually, Germany did increase testing capacities, but this came too late to prevent the significant increase in the growth rate of infections. Governments should also improve digital tracing capabilities both within their territories but more importantly across borders if they wish to avoid travel restrictions.^25^ Germany in principle has a good tracing system being built on local infrastructure but the best tracing system cannot operate if infected individuals cannot recall with whom they had close contact during their holidays.^26^ With holiday-related travel, it becomes essential that public health authorities can trace the close contacts infected people have had with those they normally do not socially interact with.

## Data Availability

Replication data will be published on dataverse.com.

https://dataverse.harvard.edu/dataverse/neumayer

